# Transdiagnostic Polygenic Risk Models for Psychopathology and Comorbidity: Cross-Ancestry Analysis in the *All of Us* Research Program

**DOI:** 10.1101/2025.03.26.25324720

**Authors:** Phil H. Lee, Jae-Yoon Jung, Brandon T. Sanzo, Rui Duan, Tian Ge, 23andMe Research Team, Irwin D. Waldman, Jordan W. Smoller, Ted Schwaba, Elliot M. Tucker-Drob, Andrew D. Grotzinger

## Abstract

Psychiatric disorders share substantial genetic liability, but the predictive utility of transdiagnostic polygenic risk scores (PRSs) remains unclear. In 102,091 *All of Us* Research Program participants, we compared a common psychiatric genetic (CPG) factor PRS with standard single-disorder and CPG-residualized disorder-specific PRSs across 11 psychiatric conditions and comorbidity burden. The CPG PRS showed stronger and more consistent associations with psychiatric diagnoses than most single-disorder-based scores, explaining up to 24.6-fold more phenotypic variance, and was strongly associated with comorbidity burden. CPG-residualized disorder-specific PRSs retained smaller independent contributions for several disorders, supporting complementary shared and disorder-specific genetic risk. Cross-ancestry analyses showed more consistent portability for comorbidity burden than for individual disorder prediction, while still underscoring limitations of Eurocentric discovery GWASs. These findings support the value of transdiagnostic PRSs as pragmatic indices of broad psychiatric liability and comorbidity burden, while emphasizing the limited utility of current PRS models as disorder-specific diagnostic tools.

## INTRODUCTION

Classification of psychiatric disorders remains one of medicine’s most challenging puzzles.^1^ While Emil Kraepelin’s foundational work established categorical diagnostic systems,^2^ clinical reality reveals overlapping symptoms and frequent comorbidities, complicating both diagnosis and treatment.^3^ This raises a fundamental question: should we conceptualize psychiatric disorders as discrete entities, or as interrelated manifestations of broader dimensions of psychopathology? For example, the Hierarchical Taxonomy of Psychopathology (HiTOP)^4,5^ and Research Domain Criteria (RDoC)^6,7^ propose that psychiatric disorders exist along continuous dimensions rather than discrete categories, better capturing shared and comorbid symptomatology.

Recent advances in psychiatric genomics have provided new tools to examine this question through the lens of inherited risk. Large-scale genomic studies have revealed extensive genetic overlap among disorders, suggesting shared biological underpinnings.^8–11^ Genomic Structural Equation Modeling (GenomicSEM)^12^ enables quantification of this shared risk through transdiagnostic polygenic risk scores (PRSs), which capture genetic vulnerability shared across multiple disorders. However, the practical utility of transdiagnostic PRSs remains uncertain. Do they improve risk stratification compared to standard single-disorder PRSs? Can they help explain psychiatric comorbidity? Does the intrinsic nature of transdiagnostic PRSs make them more generalizable across diverse populations, especially given the Eurocentric bias in psychiatric genetic research?

In this study, we address these questions by employing a common psychiatric genetic factor (CPG) approach to derive a transdiagnostic PRS designed to capture shared genetic risk across psychiatric disorders. The CPG model serves as a conceptual framework for comparing broad transdiagnostic genetic risk with disorder-specific PRSs. While our main analyses primarily utilize the CPG model, we also compare its performance with alternative transdiagnostic approaches of varying complexity^10,13^ as a secondary analysis to provide a comparative assessment of transdiagnostic risk models.

To conduct these analyses, we leverage data from the *All of Us* Research Program,^14,15^ one of the largest and most ancestrally diverse genomic initiatives to date. By integrating comprehensive health survey data on psychiatric diagnoses with participants’ genetic data, *All of Us* provides an unprecedented opportunity to examine how well transdiagnostic PRSs explain phenotypic variance in psychiatric burden and comorbidity across diagnostic categories and diverse ancestries. Specifically, we investigate the:

1. Comparative performance of CPG- versus disorder-specific PRS models across diagnostic categories.
2. Association of CPG PRSs with comorbidity burden, measured by the number of concurrent diagnoses.
3. Ancestry-related differences in predictive performance and implications for equitable risk stratification.

We hypothesize that the CPG PRS will enhance prediction of broad psychiatric burden and comorbidity by directly targeting genetic liability shared across psychiatric disorders.^16^ Under current GWAS sample-size limitations, this shared component may be estimated more precisely by aggregating genetic associations across multiple genetically correlated psychiatric GWASs. Furthermore, given known limitations of PRS generalizability derived from largely European-ancestry discovery GWASs,^17,18^ we anticipate ancestry-related attenuation of predictive performance outside European-like ancestry groups, and evaluate whether the CPG PRS shows relatively greater portability than disorder-specific PRSs. By addressing these questions, we aim to clarify the pragmatic utility of transdiagnostic PRSs for indexing broad psychiatric liability and comorbidity burden, and to contribute to the development of more equitable approaches in psychiatric genomics.

## RESULTS

### Prevalence of Psychiatric Disorders and Comorbidity

Our analysis included 102,091 individuals from the *All of Us* Research Program who completed comprehensive health surveys for psychiatric disorders (**Fig. 1**). Of these participants, 30,962 participants (30.33%) reported receiving clinical care for at least one of 13 psychiatric conditions (**Table 1**).

**Figure 1.**
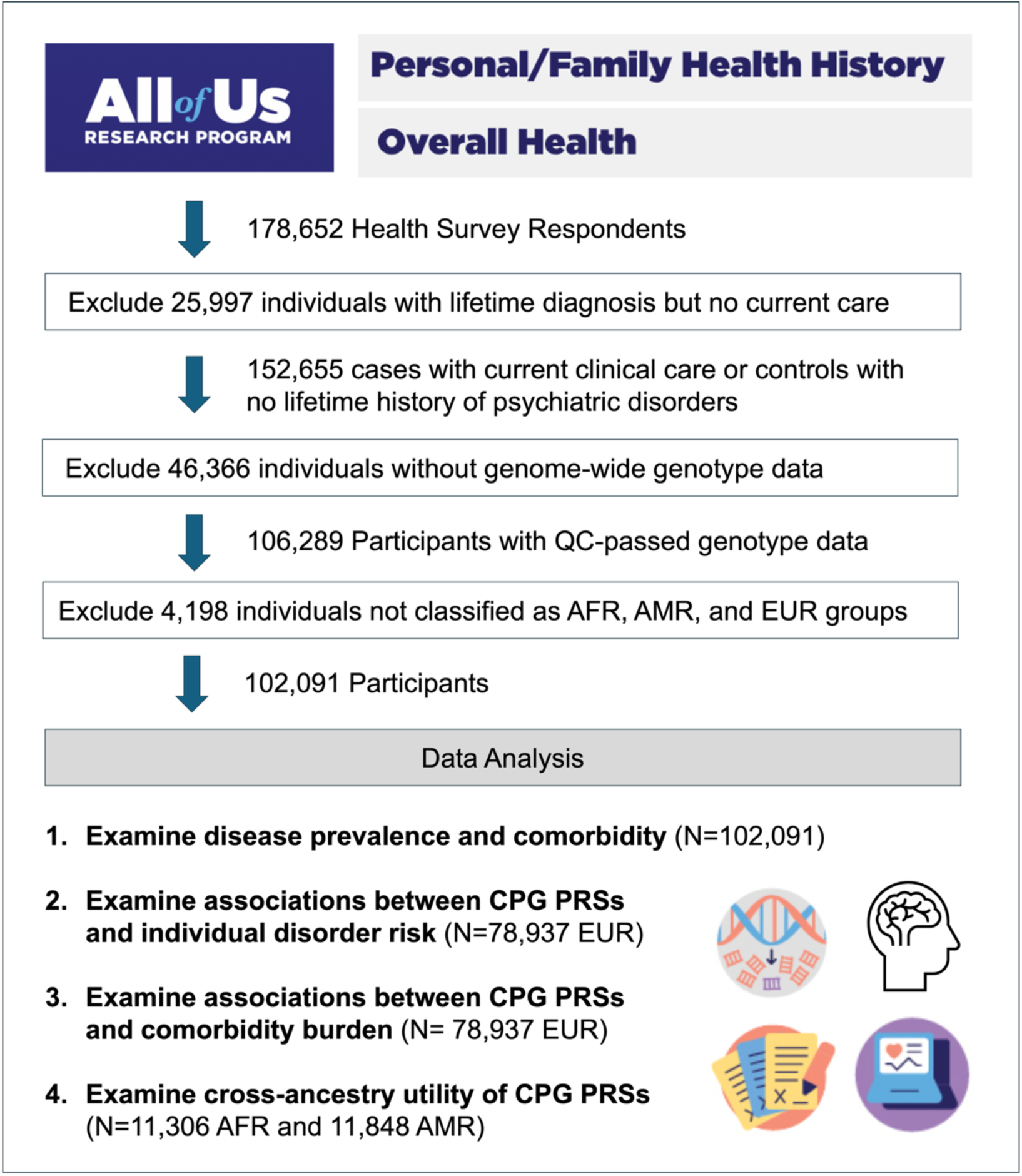
Data preparation and analysis workflow for the investigations of the common psychiatric genetic factor (CPG), psychiatric disorder risk, and comorbidity burden. The analysis cohort included a total of 102,091 participants, representing AFR (African/African American), AMR (Hispanic/Latin American), and EUR (European) ancestry groups. Given that the GWAS datasets used to generate polygenic risk scores (PRSs) were derived from EUR populations, our primary analysis focused on EUR participants (n=78,937), with separate cross-ancestry analyses conducted for AFR (n=11,306) and AMR (n=11,848) ancestry groups.

**Table 1.** Prevalence of 13 Psychiatric Conditions Assessed in the *All of Us* Personal and Family Health Survey (N=102,091). Participants were classified as cases if they reported both a diagnosis and current clinical care for a given condition. Sample prevalence represents the proportion of participants meeting these criteria within the study sample. Population prevalence (12-month) estimates are provided for reference from external epidemiological studies. “-” indicates that a population prevalence estimate was not available.

| **Surveyed Conditions** | **Case Number** | **Sample Prevalence (%)** | **Population Prevalence (12 month) (%)** |
| --- | --- | --- | --- |
| Depression | 22,546 | 22.08 | 8.3 |
| Anxiety Reaction/Panic Disorder | 17,451 | 17.09 | 2.7 |
| Post-Traumatic Stress Disorder | 5,590 | 5.48 | 3.6 |
| Attention-Deficit Hyperactivity Disorder | 3,899 | 3.82 | 4.4 |
| Bipolar Disorder | 3,349 | 3.28 | 2.8 |
| Social Phobia | 1,350 | 1.32 | 7.1 |
| Alcohol Use Disorder | 1,197 | 1.17 | 4.4 |
| Eating Disorder | 1,152 | 1.13 | 1.2 |
| Other Mental Health or Substance Use Condition | 998 | 0.98 | - |
| Personality Disorder | 868 | 0.85 | 1.4 |
| Drug Use Disorder | 809 | 0.79 | 3.9 |
| Schizophrenia | 449 | 0.44 | 0.6 |
| Autism Spectrum Disorder | 273 | 0.27 | 0.6 |

Mood and anxiety disorders were the most prevalent, with depression affecting 22.08% and anxiety reaction/panic disorders affecting 17.09% of the cohort. Other conditions with notable prevalence rates included post-traumatic stress disorder (PTSD) at 5.48%, attention-deficits/hyperactivity disorder (ADHD) at 3.82%, and bipolar disorder at 3.28%. Autism spectrum disorders exhibited the lowest prevalence at 0.27%.

Significant levels of comorbidity were evident across psychiatric disorders, as indicated by tetrachoric correlations ranging from modest (r = 0.16) to very strong (r = 0.78) (**Supplementary Table 1**). The strongest comorbid relationships were identified between anxiety reaction/panic disorder and depression (r = 0.78) and between drug use disorder and alcohol use disorder (r = 0.72). Depression also showed strong correlations with PTSD (r = 0.67), personality disorder (r = 0.65), and bipolar disorder (r = 0.59). Conversely, autism spectrum disorders demonstrated the weakest correlations overall, particularly with substance use disorders (r < 0.2).

Among participants with any psychiatric condition, 53% (16,539/30,962) reported multiple (≥ 2) disorders requiring current clinical care (**Supplementary Table 2**). These findings underscore the pervasive nature of comorbidity, reinforcing the need for integrative approaches in diagnosis, treatment planning, and risk prediction.

### Associations of the CPG and Disorder-Specific PRSs with Psychiatric Disorders

Of the 13 psychiatric conditions surveyed in the *All of Us* Program (**Table 1**), GWAS data were closely matched for 11 disorders, allowing for direct comparisons of disorder-specific and CPG genetic risks. These included depression, ADHD, alcohol use disorder, anxiety reaction/panic disorder, autism spectrum disorder, bipolar disorder, drug use disorder, eating disorder, PTSD, schizophrenia, and social phobia. For these 11 disorders, we assessed associations with three types of PRSs in European ancestry participants (*n* = 78,937): (1) the CPG PRS, representing shared genetic risk across 15 psychiatric disorders calculated using the multivariate GWAS summary statistics from GenomicSEM^12^; (2) standard single-disorder PRSs, calculated from GWAS summary statistics for each individual disorder and reflecting both shared and disorder-indexed genetic liability; and (3) CPG-residualized disorder-specific PRSs, representing genetic effects unique to each disorder not explained by the CPG PRS (see **Methods, Supplementary Tables 3-5**).

Standard univariate disorder PRSs were significantly associated with their corresponding disorders (all FDR p < 0.001, **Fig. 2A, Supplementary Table 6**). The CPG PRS also showed significant positive associations with all 11 psychiatric disorders (all FDR p < 0.001). Moreover, the CPG PRS demonstrated significantly larger effect sizes compared to both standard single-disorder PRSs and CPG-residualized disorder-specific PRSs: the median odds ratios (OR) for CPG PRSs were 1.58, compared to 1.42 for standard disorder PRSs (two-sided Wilcoxon rank-sum test *p*=1.14×10^-2^) and 1.17 for disorder-specific PRSs (two-sided Wilcoxon rank-sum test *p*=1.42×10^-5^).

**Figure 2.**
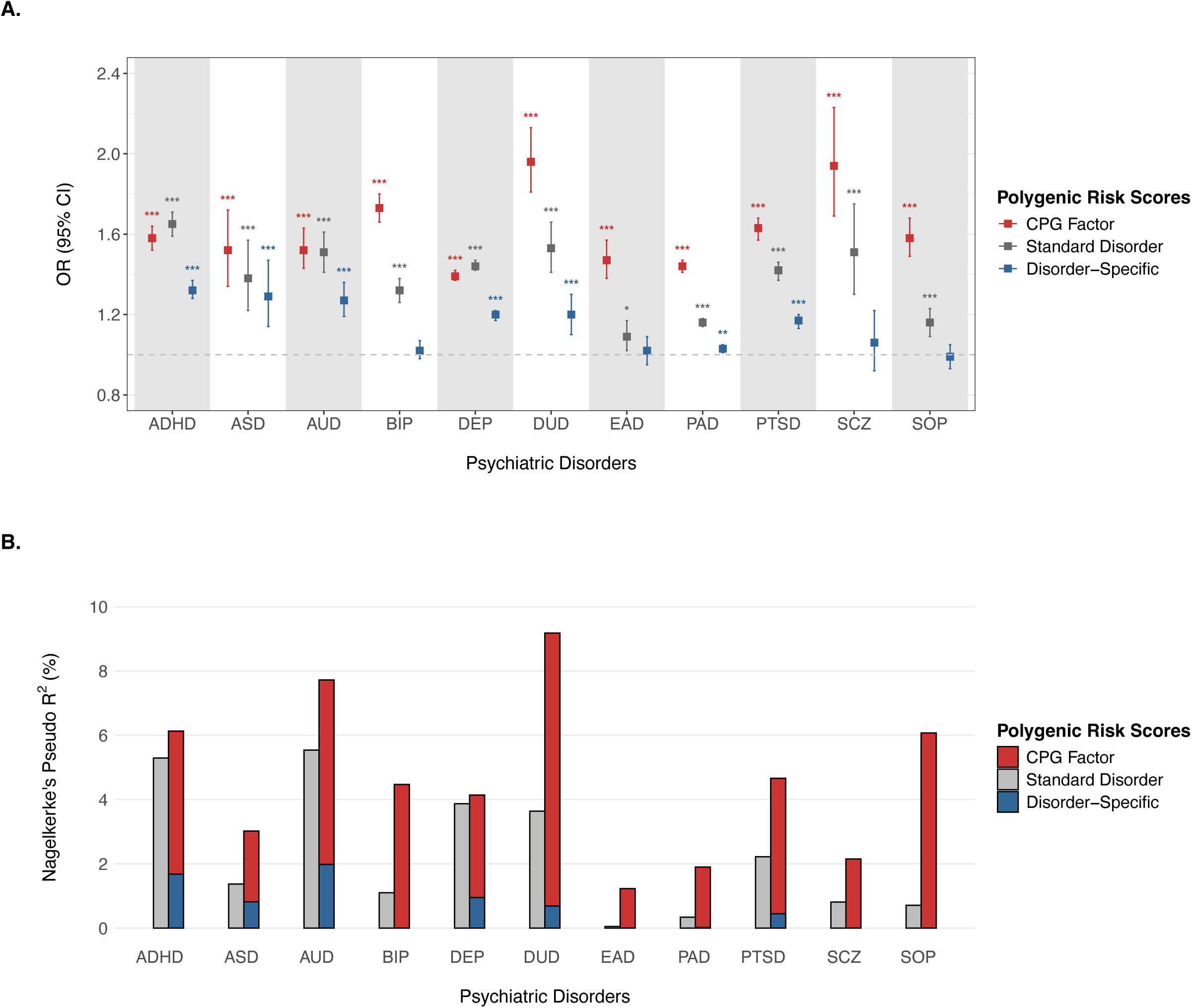
Association findings of the common psychiatric genetic factor (CPG) and disorder-targeted polygenic risk scores (PRSs) across 11 psychiatric disorders in the European ancestry population. A. Odds ratios (OR) plot. The forest plot displays OR with 95% confidence intervals (CI) for associations between psychiatric disorders and three types of polygenic risk scores (PRS): standard disorder PRS (gray), disorder-specific PRS (dark blue), and CPG PRS (red). Significance levels are indicated by asterisks: * FDR (false discovery rate) p < 0.05, ** p < 0.01, *** p < 0.001. B. Phenotypic variance explained by PRSs. The y-axis shows liability-based Nagelkerke’s pseudo *R²* values (%) for three PRS categories: standard disorder PRS (gray), disorder-specific PRS (dark blue), and CPG PRS (red). For each disorder, individual PRS effects are shown in the left bar, while the stacked bars on the right display the additive effects of the shared (CPG) and disorder-specific PRS components. Disorders are arranged by alphabetical orders. ADHD: attention-deficit/hyperactivity disorder; ASD: autism spectrum disorder; AUD: alcohol use disorder; BPD: bipolar disorder; EAD: eating disorder; PAD: anxiety reaction/ panic disorder; PTSD: post-traumatic stress disorder; SCZ: schizophrenia; SOP: social phobia.

For six disorders, ADHD, autism spectrum disorder, alcohol use disorder, depression, drug use disorder, and PTSD, both shared genetic risk (captured by the CPG PRS) and disorder-specific genetic risk made significant independent contributions. For instance, the CPG PRS for ADHD had an OR of 1.58 (95% CI: 1.52–1.64), while the disorder-specific ADHD PRS independently retained an OR of 1.32 (95% CI: 1.28–1.37). Similarly, for autism spectrum disorder, the CPG PRS showed an OR of 1.52 (95% CI: 1.34–1.72), and the disorder-specific ASD PRS exhibited an independent OR of 1.29 (95% CI: 1.14–1.47).

When evaluating phenotypic variance (R²) explained by these genetic scores, the CPG PRS consistently outperformed residualized disorder-specific PRSs across all psychiatric conditions (two-sided Wilcoxon rank-sum test *p*=2.45×10^-3^). **Fig. 2B** demonstrates that incorporating both the CPG and CPG-residualized disorder-specific PRSs provides additional predictive power beyond standard single-disorder PRSs alone (**Supplementary Table 7**). This improvement was particularly pronounced for eating disorder (increasing variance explained from 0.05% to 1.23%, a 24.6-fold increase), social phobia (from 0.71% to 6.07%, 8.5-fold), drug use disorder (from 3.64% to 9.18%, 2.5-fold), and bipolar disorder (from 1.10% to 4.47%, 4.1-fold), where standard single-disorder PRSs explained limited phenotypic variance.

To ensure robustness, sensitivity analyses were conducted by varying diagnostic criteria and control group definitions. Both lifetime and current diagnoses based on medication produced consistent findings (**Supplementary Tables 8,9**). In contrast, analyses of lifetime diagnoses without current clinical care showed weaker genetic associations compared to those under current care, which could reflect either attenuation due to recall bias or differences in the persistence of psychiatric conditions over time (**Supplementary Table 10**). Additionally, analyses utilizing control groups with no corresponding disorder with current clinical care yielded consistently significant findings, albeit with modestly smaller PRS effects than our primary findings (one-sided pairwise t-test *p* = 6.72×10^-8^; **Supplementary Tables 11**).

### Associations of the CPG and Disorder-Specific PRSs with Psychiatric Comorbidity

We next examined the associations between PRSs and the burden of psychiatric comorbidity, measured by the number of concurrent psychiatric disorders under clinical care (**Fig. 3, Supplementary Table 12**). The CPG PRS demonstrated a strong association with comorbidity burden (standardized beta coefficient β = 0.26, SE = 0.006, *R*^2^ = 3.41%, FDR p < 0.05). Among standard single-disorder PRSs, all except the obsessive-compulsive disorder PRS were significantly associated with psychiatric comorbidity. The depression PRS exhibited the strongest effect (β = 0.263, SE = 0.006, *R*^2^ = 3.53%), followed by ADHD (β = 0.202, SE = 0.006, *R²* = 2.1%) and PTSD (β = 0.146, SE = 0.006, *R²* = 1.09%).

**Figure 3.**
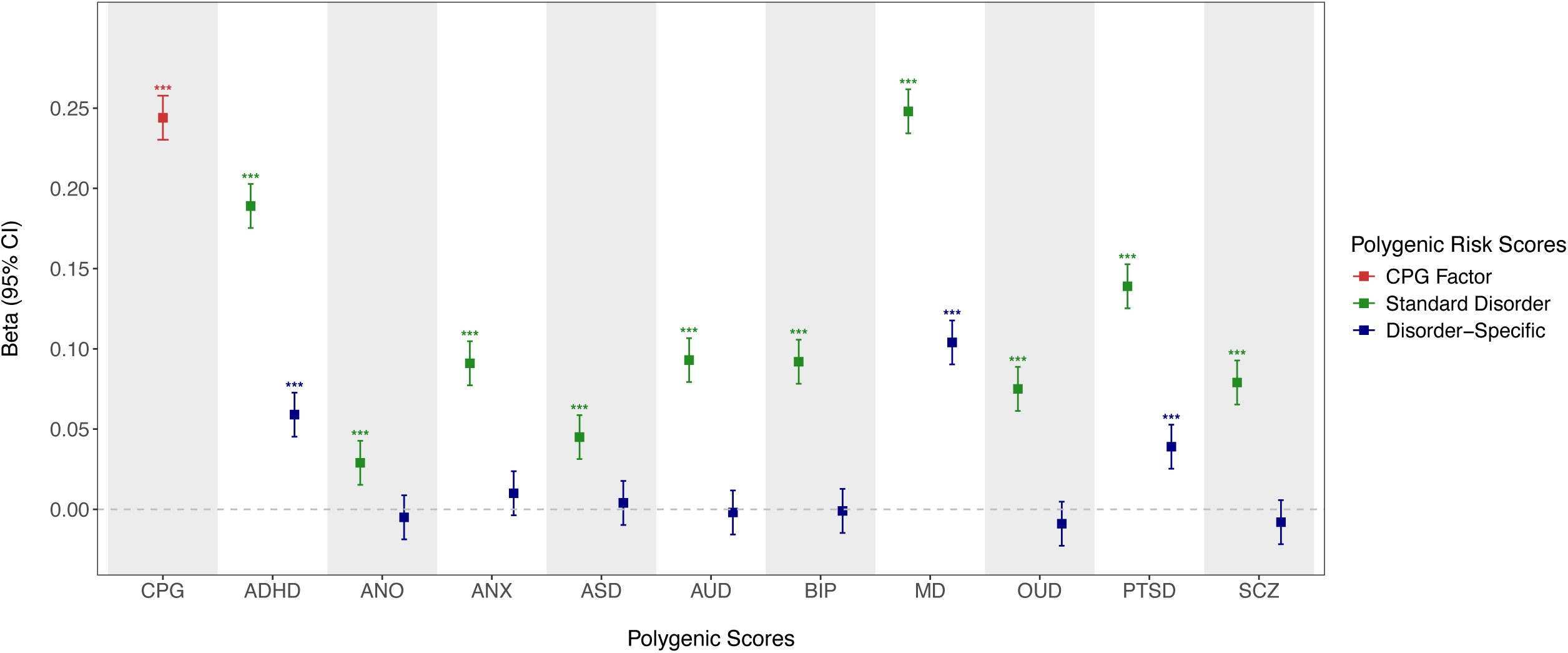
Differential associations of the common psychiatric genetic factor (CPG) and disorder-specific polygenic risk scores (PRSs) with psychiatric comorbidity in the European ancestry population. The plot displays beta coefficients and with 95% confidence intervals (CI) for associations between PRSs and psychiatric comorbidity. Three types of PRS are compared: standard disorder PRS (gray), disorder-specific PRS with CPG component removed (navy), and CPG PRS (red). The x-axis shows different psychiatric disorder PRSs: MD (major depression), ADHD (attention-deficit/hyperactivity disorder), PTSD (post-traumatic stress disorder), AUD (alcohol use disorder), BIP (bipolar disorder), ANX (anxiety), SCZ (schizophrenia), OUD (opioid use disorder), CUD (cannabis use disorder), NID (nicotine dependence), ASD (autism spectrum disorder), CAD (cocaine addiction), TS (Tourette syndrome), ANO (anorexia nervosa), and OCD (obsessive-compulsive disorder). Significance levels are indicated by asterisks: * FDR (false discovery rate) p < 0.05, ** p < 0.01, *** p < 0.001.

When the CPG component was removed, most disorder-specific effects were substantially attenuated. Three disorders, however, retained significant positive disorder-specific effects, with independent contributions to variance explained (*R*^2^): depression (β = 0.11, SE = 0.006, *R*^2^ = 0.63%), ADHD (β = 0.063, SE = 0.006, *R*^2^ =0.21%), and PTSD (β = 0.039, SE = 0.006, *R*^2^ =0.08%). These significant disorder-specific contributions likely reflect both the high prevalence of these conditions in the cohort (**Table 1**) and the large, well-powered GWAS datasets used to derive their PRSs (**Supplementary Table 3).** Taken together with the CPG PRS, these PRSs explained a total of 4.38% of the variance in psychiatric comorbidity (**Supplementary Table 12**).

### Cross-Ancestry Evaluation of the CPG and Disorder-Specific PRSs

We examined the performance of the CPG and disorder-specific PRSs in predicting individual psychiatric conditions and comorbidity burden across AFR- and AMR-like participants, comparing these results to the EUR cohort presented earlier (**Fig. 4, Supplementary Tables 13-14**).

**Figure 4.**
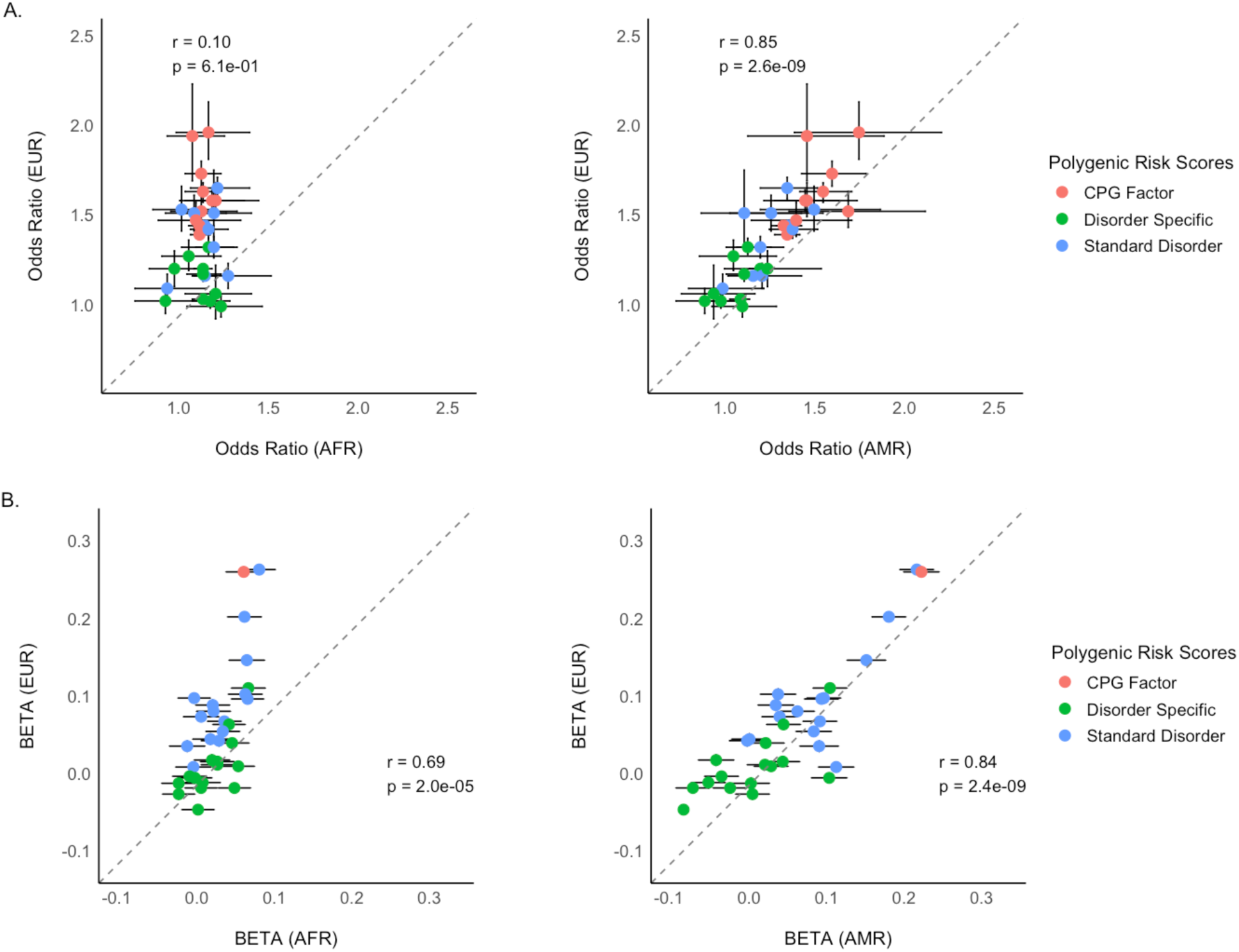
Cross-ancestry applicability of European-derived polygenic risk scores (PRSs) in African/African American (AFR) and Hispanic/Latin American (AMR) ancestry groups. **A.** Odds ratio (OR) plot showing the associations between PRSs (CPG factor, disorder-specific, and standard disorder PRSs) and 11 psychiatric disorders surveyed in the *All of Us* data. ORs for each ancestry group are compared against those derived from the European (EUR) cohort. The dotted line represents the unity line, with correlations (r) and significance (p-values) shown for AFR and AMR relative to EUR. **B.** Regression beta plot assessing the associations between the CPG and 15 psychiatric disorder PRSs and psychiatric comorbidity burden, measured as the number of concurrent psychiatric conditions. Effect sizes (β coefficients) for AFR and AMR groups are compared with those from the EUR cohort. The CPG PRS showed among the strongest and most consistent associations with comorbidity burden across ancestry groups, with significant cross-ancestry correlations observed for both AFR- and AMR-like participants.

For individual disorder prediction (**Fig. 4A, Supplementary Table 13**), PRS effects were weakest in the AFR group, with few significant associations for the CPG PRS or disorder-specific PRSs, and the ORs showing minimal correlations with those observed in EUR (r = 0.10, *p* = 6.13 × 10⁻¹). In contrast, PRS performance was more consistent in AMR, with highly correlated ORs for both CPG and CPG-residualized disorder-specific PRSs (r = 0.85, p = 2.57 × 10⁻⁹).

When examining comorbidity burden (**Fig. 4B, Supplementary Table 14**), the CPG PRS showed significant associations in all three ancestry groups, including AFR, and effect sizes (β coefficients) were significantly correlated between AFR and EUR (r = 0.69, p = 2.0 × 10⁻⁵). Correlations were stronger for AMR, with β coefficients closely aligned with EUR (r = 0.84, p = 2.4 × 10⁻⁹). Across all ancestry groups, the CPG PRS explained more variance in psychiatric risk (mixed effects β=0.28, SE=0.09, p-value=3.78×10^-3^) and comorbidity burden (mixed effects β=1.48, SE=0.30, p-value=4.28×10^-6^) than CPG-residualized disorder-specific PRSs, indicating improved cross-ancestry transferability.

### Comparison with Alternative Transdiagnostic Models

We examined whether more sophisticated transdiagnostic PRS models offer predictive advantages beyond the parsimonious CPG approach. We compared the CPG model with correlated four- and five-factor models building on the work by Grotzinger et al.^10,13^ The four-factor model categorized 11 psychiatric disorders into compulsive/obsessive (F1), thought (F2), neurodevelopmental (F3), and internalizing (F4) disorders^10^, while we further extended this framework to include a substance disorder (F5) dimension across the same 15 disorders^19–42^ used to define the CPG (see **Methods**, detailed modeling results in **Supplementary Table 15**).

Despite the superior model fit of multifactor models in GenomicSEM (CFI=0.96-0.97 vs 0.77), the simpler CPG model demonstrated comparable or superior predictive performance (**Fig. 5, Supplementary Tables 16-17**). The neurodevelopmental disorder factor (F3) showed the strongest performance for ADHD among the factor-specific PRSs, though still not surpassing CPG. Similarly, the substance-use disorder factor (F5) showed comparable performance to CPG for alcohol use disorder, supporting the specificity of these latent factors. Notably, the internalizing factor (F4) demonstrated performance comparable to CPG across multiple disorders, exhibiting its broad transdiagnostic effects.

**Figure 5.**
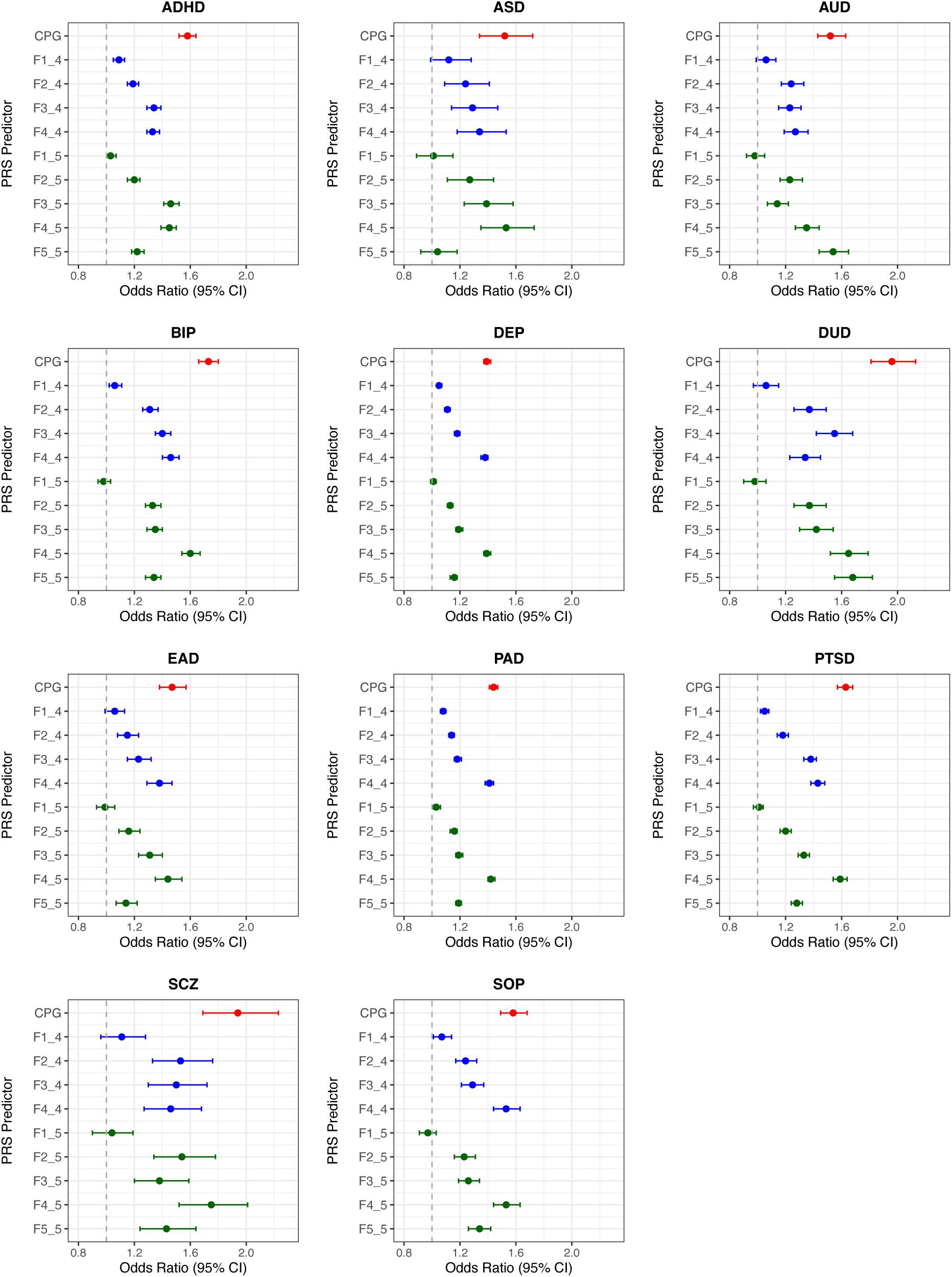
Comparison of PRSs based on the common psychiatric genetic factor (CPG) and correlated multifactor models across 11 psychiatric conditions. The figure consists of 11 forest plots, each representing one of 11 psychiatric disorders: ADHD, ANX/PD (anxiety reaction/panic disorder), ASD (autism spectrum disorder), AUD (alcohol use disorder), BIP (bipolar disorder), DEP (depression), DUD (drug use disorder), ED (eating disorder), PTSD (post-traumatic stress disorder), SCZ (schizophrenia), and SOP (social phobia). Each plot displays odds ratios (OR) with 95% confidence intervals for different PRS predictors. The predictors include the Common Psychiatric Genomic PRS (CPG, shown in red), four correlated-factor model PRSs (F1_4 through F4_4, shown in blue), and five correlated-factor model PRSs (F1_5 through F5_5, shown in green). The four correlated factor model, introduced by Grotzinger et al. (2022)^10^, represent: F1_4: compulsive/obsessive, F2_4: thought disorder, F3_4: neurodevelopmental disorder, and F4_4: internalizing disorder. The five correlated factor model, estimated in this study (Supplementary Table 15), represent: F1_5: compulsive/obsessive, F2_5: schizophrenia/bipolar disorder, F3_5: neurodevelopmental disorder, F4_5: internalizing disorder, and F5_5: substance use disorder. The x-axis represents OR ranging from 0.8 to 2.3, with a vertical line at 1.0 indicating no effect. Data points to the right of this line represent positive associations between genetic risk scores and disorder prevalence.

## DISCUSSION

In this study, we leveraged data from the *All of Us* Research Program,^14,15^ a large and demographically diverse US cohort, to evaluate transdiagnostic and disorder-specific polygenic risk models for psychiatric disorders. By integrating comprehensive health survey data with genome-wide genetic data, we demonstrate that the CPG PRS serves as a robust index for both individual psychiatric diagnoses and the overall burden of comorbidity. Reflecting shared genetic risk across 15 psychiatric disorders, the CPG PRS generally matched or outperformed standard single disorder-based PRSs across conditions. This improvement was particularly evident for conditions like social phobia, where variance explained increased from 0.71% to 6.07% (8.5-fold increase), demonstrating how the CPG model can recover significant predictive signal for disorders that are currently under-powered in single-disorder GWAS.

The strong predictive performance of the CPG risk model may be driven by several interrelated factors. First, the CPG may capture transdiagnostic genetic liability that cut across broad diagnostic categories. Prior cross-disorder and pleiotropy studies suggest that this shared liability may refelct fundamental neurobiological mechanisms, such as transcription regulation and early neurodevelopmental pathways.^11^ At the phenotypic level, this shared liability may also align with transdiagnostic biobehavioral dimensions, such as emotion sensitivity or distress tolerance.^11^ Second, the CPG PRS benefits from significantly enhanced statistical power by aggregating data across multiple genetically correlated GWAS datasets. This aggregation combines numerous small effects into a more reliable measure of genetic liability, effectively reducing the noise inherent in single-disorder GWAS.

Finally, the phenotypic complexity of the *All of Us* cohort likely prioritize transdiagnostic signals. While discovery GWASs are typically designed to isolate single-disorder-targetted etiological pathways, our sample exhibited pervasive comorbidity, with 53% of affected individuals reporting multiple concurrent diagnoses. In such a population, shared genetic risk (captured by the CPG PRS) may serve as a more robust and pragmatic index of broad psychiatric liability than disorder-specific residuals targeting unique, non-overlapping variance.

The relevance of the CPG PRS was further supported by its robust association with psychiatric comorbidity burden. Individuals with higher CPG scores were more likely to report multiple psychiatric diagnoses, reinforcing the relevance of shared genetic liability for understanding the high comorbidity observed in clinical and population settings. While standard PRSs for depression, ADHD, and PTSD also showed associations with comorbidity burden, these effects were substantially attenuated after removal of the CPG component, indicating that much of their association with comorbidity was attributable to shared transdiagnostic risk.

Nevertheless, the stronger performance of the CPG PRS in this cohort does not diminish the importance of disorder-specific genetic risk. As discussed above, the predictive advantage of the CPG PRS likely reflects a combination of true shared genetic liability, increased discovery power, and the highly comorbid phenotypic structure of the *All of Us* cohort, which cannot be clearly separated under current data constraints. Indeed, CPG-residualized disorder-specific PRSs retained significant independent contributions for several disorders, including ADHD, autism spectrum disorder, alcohol use disorder, and depression. These findings indicate that individual disorders retain distinct genetic liability not fully captured by a purely transdiagnostic approach. Conversely, weak or null residualized effects for certain disorders, such as schizophrenia and bipolar disorder, should not be interpreted as evidence that these disorders lack disorder-specific genetic influences. Residualized PRS analyses are sensitive to target-sample case counts, phenotype definition, phenotypic comorbidity, discovery-GWAS precision, and the amount of disorder-specific genetic signal remaining after shared transdiagnostic liability has been accounted for. Thus, the CPG PRS is best viewed as a pragmatic index of broad psychiatric liability, while disorder-specific PRSs provide complementary, although currently less consistently detected, information about residual diagnostic risk. As discovery GWAS sample sizes continue to grow, we anticipate that the precision of disorder-specific risk estimates will improve, allowing future studies to better isolate genetic liability that is more specific to individual psychiatric conditions.^43–45^

When benchmarked against more differentiated transdiagnostic models, the CPG PRS remained a strong and parsimonious predictor. This comparison illustrates the distinction between explanatory and predictive frameworks.^46^ While correlated multifactor models provided a more detailed representation of the complex genetic architecture underlying psychiatric disorders,^8–11^ the simpler CPG PRS performed comparably to, or better than factor-specific PRSs for several predictive outcomes. Importantly, this finding should not be interpreted as evidence that psychiatric genetic architecture is best represented by a single common factor. Rather, the CPG PRS provides a pragmatic predictive approach by efficiently leveraging shared genetic signal across disorders, whereas multifactorial and hierarchical models remain essential for understanding etiological structure. More systematic benchmarking across transdiagnostic PRS frameworks will help clarify their relative strengths and limitations, including when more differentiated dimensions of genetic liability provide incremental predictive value.

Finally, beyond model comparisons, our findings highlight a nuanced perspective on the cross-ancestry portability of psychiatric PRSs.^17,47^ While individual-disorder prediction remained limited outside European-like ancestry groups, PRS associations with overall comorbidity burden showed more consistent cross-ancestry patterns, with effect-size correlations of r = 0.69 for AFR and r = 0.84 for AMR relative to EUR. Under current EUR-derived PRS models, broad psychiatric burden may provide a more stable target for cross-ancestry prediction than specific psychiatric diagnoses.

Several limitations warrant consideration. First, the *All of Us* cohort is US-based, which may limit global generalizability. Second, grouping individuals into broad ancestry groups (AFR, AMR, and EUR) may oversimplify the nuanced and heterogeneous nature of human genetic variation. Third, psychiatric outcomes were based on self-reported diagnoses, which may introduce misclassification and recall bias.^48^ Structured diagnostic interviews or EHR-validated phenotypes would further strengthen future studies. Fourth, null or weak residualized disorder-specific effects should be interpreted cautiously and may reflect substantial overlap with shared transdiagnostic liability, limited power to detect residual effects, phenotype heterogeneity, and discovery–target phenotype differences, rather than absence of disorder-specific genetic risk. Thus, our results should be interpreted as predictive comparisons under currently available discovery GWASs, rather than as a definitive test of whether transdiagnostic or disorder-specific PRSs would perform better under equal discovery power. Lastly, the predictive performance of current PRSs remains modest and inadequate for clinical use,^49^ especially in non-European populations.

In conclusion, our study supports an integrated predictive framework that combines transdiagnostic and disorder-specific genetic risk. The transdiagnostic CPG PRS is particularly informative for broad psychiatric risk and comorbidity burden, while CPG-residualized disorder-specific PRSs add complementary, though generally smaller, risk information. Future studies with more diverse genomic datasets, improved psychiatric phenotyping, and advanced PRS methods that jointly model transdiagnostic and residualized disorder-specific genetic liability will be essential to clarify how these complementary sources of risk can be optimally combined to improve future risk-assessment frameworks and inform precision psychiatry research.

## ONLINE METHODS

### *All of Us* Research Program

The *All of Us* Research Program^14^ is a nationwide, longitudinal study aimed at advancing precision medicine by recruiting a diverse cohort of participants across the U.S. The study collects extensive health-related data, including genetic information, electronic health records, lifestyle factors, medical history surveys, and environmental exposures. Our analysis utilizes the curated data v7 (2022Q4R9), which includes enrollment participant data up to July 1, 2022. The present study was performed between November 2023 and February 2025 using the *All of Us* Researcher Workbench, a cloud-based platform where approved researchers can analyze *All of Us* data.

### Outcome Measures

Psychiatric disorder diagnoses were obtained from the *All of Us Personal and Family Health History* survey (https://www.researchallofus.org/data-tools/survey-explorer/). Participants self-reported whether they had been diagnosed with each of the following mental health and substance use conditions: alcohol use disorder, anxiety reaction/panic disorder, attention-deficit/hyperactivity disorder (ADHD), autism spectrum disorder, bipolar disorder, depression, drug use disorder, eating disorder, personality disorder, post-traumatic stress disorder (PTSD), schizophrenia, social phobia, and other mental health conditions. To improve case validity, participants were classified as cases if they reported both a diagnosis and current clinical care (affirmative response to “*still seeing a doctor or health care provider?*”). Controls were those with no psychiatric disorder diagnoses.

We also computed an overall measure of concurrent psychiatric comorbidity as the total number of the aforementioned psychiatric disorders for each participant. These cumulative counts ranged from 0 (no psychiatric disorder) to 13 (participants with all surveyed conditions).

### Genetic Ancestry Assignment

Out of 178,652 Health Survey respondents, 106,289 participants had QC-passed genome-wide genetic data with case/control phenotype assignment (**Figure 1**). Genetic ancestry groups were defined using the *All of Us*-provided predicted genetic ancestry labels derived from genomic data, rather than self-reported race or ethnicity. The predicted ancestry labels and ancestry probabilities for major reference-like groups included African/African American-like (AFR), admixed American-like (AMR), East Asian-like (EAS), European-like (EUR), Middle Eastern-like (MID), and South Asian-like (SAS), with an additional “Other” category for samples not confidently assigned to one of these groups. In the present study, analyses were restricted to 102,091 participants assigned to EUR- (N=78,937), AFR- (N=11,306), or AMR-like (N=11,848) genetic ancestry groups because these groups had sufficient overall sample size and diagnosis-specific case counts to support stable regression estimates across the 11 psychiatric outcomes examined. Participants assigned to EAS-, MID-, SAS-like, or Other groups were not analyzed separately because sample sizes and case counts for several psychiatric outcomes were too small for stable estimation and disclosure-compliant reporting.

### Common Genetic Factor of Psychopathology (CPG)

To model shared genetic variance across psychiatric disorders, we assembled GWAS datasets for 15 psychiatric disorders: ADHD, alcohol use disorder, anorexia nervosa, anxiety disorder, autism spectrum disorder, bipolar disorder, major depression, PTSD, cannabis use disorder, cocaine addiction, nicotine dependence, obsessive compulsive disorder, opioid use disorder, Tourette’s syndrome, schizophrenia (**Supplementary Table 3**). These datasets, derived from European-ancestry populations due to limited representation of other ancestries, included conditions directly assessed in the *All of Us* survey data (e.g., ADHD, alcohol use disorder, schizophrenia, bipolar disorder) or closely aligned with survey-derived measures (e.g., substance use disorders (e.g., cannabis, cocaine, nicotine, and opioid) for drug use disorder). The inclusion of additional GWAS datasets was intended to enhance the robustness and generalizability of the common psychiatric genetic factor.

We fitted a common factor model in GenomicSEM^12^ to estimate shared genetic variance across 15 psychiatric disorders. We selected this parsimonious approach for its conceptual clarity and to facilitate direct comparisons between the transdiagnostic effects and disorder-specific effects. Further details, including disorder loadings onto the common factor, are provided in **Supplementary Table 4**.

### Polygenic Risk Score Predictors

Genotyping, imputation, and quality control (QC) procedures for the *All of Us* data have been detailed elsewhere.^15^ PRS predictors were separately calculated for participants of African/African American (AFR), admixed Hispanic/Latino American (AMR), and European descent (EUR) using PRS-CS^50^ and PLINK.^51^ Specifically, we used PRS-CS-auto, in which the global shrinkage parameter (phi) was not fixed but learned from the data using the fully Bayesian implementation. Except 10,000 total MCMC iterations and 5,000 burn-in iterations, we used the default PRS-CS hyperparameters: gamma-gamma prior parameters a = 1, b = 0.5, and a thinning factor of 5. Because the discovery GWASs used in this study were derived primarily from European-ancestry samples, we used the 1000 Genomes Project European LD reference panel, consistent with PRS-CS guidance that the LD reference panel should match the ancestry of the GWAS sample rather than the target sample.

Using the genome-wide genetic data estimated from the common factor model, we calculated PRS_CPG_ using PLINK as follows:

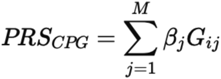

where *i* indexes the individual, *j* indexes each variant, *M* is the total number of genetic variants included in the score, the weight (β_j_) is the posterior SNP effect size obtained from PRS-CS^50^ for variant *j*, *G_ij_* is the number of effect alleles, 0,1, and 2. To remove potential population substructure, we regressed PRSs on the top ten genetic PCs as below:

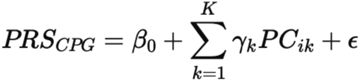

where β_0_ is the intercept, PC*_ik_* are the *K* ancestry-specific genetic PCs, γ_k_ are the regression coefficients for the k-th PC, and ɛ represents the residuals. After fitting the regression, we used the residual as the PRS_CPG_.

Similarly, we calculated two types of PRSs for each of the 15 psychiatric disorders. The standard disorder PRS for a specific disorder (e.g., PRS_SCZ_) was calculated in the same way as described previously using GWAS summary statistics specific to the corresponding disorder.

Additionally, we calculated disorder-specific PRS (e.g., PRS_SCZ-CPG_) that indexes disorder-specific genetic risk independent of the shared genetic variance captured by the common factor. We used a linear regression approach where the standard disorder PRS was modeled as a function of PRS_CPG_ and ancestry-specific genetic principal components (PCs). The residuals from this regression represent the disorder-specific PRS:

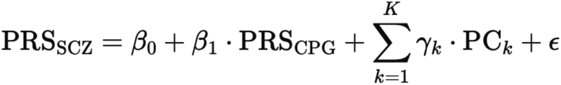

where β_0_ is the intercept, β_1_ is the regression coefficient for PRS_CPG_, PC_k_ are *K* ancestry-specific genetic PCs, γ_k_ are the regression coefficients for the k-th PC, and ɛ represents the residuals.

To account for ancestry-specific genetic architecture, derivation of genetic principal components (PCs) and PRS calculation were performed separately within each ancestry group.

### Association Analysis of PRS and Outcome Measures

For binary outcome measures (individual disorder diagnosis), we examined the associations between each PRS predictor and outcome measure using logistic regression via the *glm* function in the *lme4* R package (version 4.4.2). For comorbidity variables, which represent the count of concurrent psychiatric diagnoses, we used negative binomial regression. This approach extends Poisson regression by allowing for overdispersion, a feature noticed in our count data, and was tested using the *glm.nb* function from the MASS R package. All regression models included the following covariates: age, self-reported sex, genotyping batch, and the top ten genetic PCs to adjust for within-ancestry population structure. Continous covariates were standardized.

To evaluate the proportion of variance explained by each PRS, we calculated *Nagelkerke’s pseudo R²* for binary outcomes and adjusted R² for continuous outcomes. As observed R^2^ were not directly comparable between disorders with different sample prevalence, we calculated liability-based R^2^ as follows^52^:

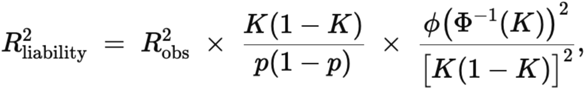

where *K* is the population prevalence, *p* is the sample prevalence, *Φ^−1^(K)* is the liability threshold for prevalence *K* under a standard normal distribution, and *ϕ* is its probability density function.

The statistical significance of variance uniquely attributable to PRS predictors was assessed using likelihood ratio tests (LRTs). LRTs compared full models (including the PRS) to null models (excluding the PRS), providing a robust measure of the incremental predictive power of PRS predictors. All p-values were corrected for multiple comparisons using the false discovery rate (FDR) to control for type I error across the large number of tests conducted.

Pairwise comparisons of odds ratios (ORs) between the three groups (CPG vs standard disorder vs. disorder-specific PRSs) were conducted using Wilcoxon rank-sum tests. Statistical significance was set at p < 0.05 after multiple testing correction. Group medians were calculated to quantify the magnitude of differences between pairs.

Given that the GWAS datasets used for PRS derivation were exclusively based on European-ancestry individuals, the primary analyses were conducted on participants of European descent to minimize potential confounding due to ancestry-specific differences in genetic architecture. To examine the cross-ancestry generalizability of PRS predictors, separate analyses were conducted on AFR and AMR participants and these results are presented in a dedicated section.

### Sensitivity Analyses

To ensure robustness, we conducted sensitivity analyses addressing diagnostic criteria, control group definitions, and comorbidity measures. Associations were tested using both lifetime and current diagnoses to evaluate the impact of temporal scope on PRS-outcome relationships. Control group heterogeneity was assessed by comparing results using individuals without current diagnoses versus those without any lifetime diagnoses. For comorbidity outcomes, winsorization was applied to cap extreme values at the 95th percentile, mitigating the influence of outliers and ensuring consistency in findings across transformations.

### Correlated Factor Models

To assess the predictive performance of the CPG risk model in comparison to more complex latent factor models, we developed a five correlated factor model using the same 15 psychiatric disorders (**Supplementary Table 3**) analyzed in our CPG approach using GenomicSEM.^12^ First, we estimated genetic correlations between all disorder pairs using LD Score Regression (LDSC).^53^ We then performed exploratory factor analysis (EFA) on the genetic correlation matrix to determine the optimal number of factors, evaluating models with 2-6 factors. Model selection was guided by comparative fit indices (CFI) and standardized root mean square residual (SRMR). The five-factor solution demonstrated superior fit compared to models with fewer factors (p-value of χ^2^ = 4.49×10^-95^; CFI = 0.97, SRMR = 0.78) while remaining parsimonious and interpretable. The resulting five correlated factors were characterized as: compulsive/obsessive disorders (F1_5), schizophrenia/bipolar disorders (F2_5), neurodevelopmental disorders (F2_5), internalizing disorders (F4_5), and substance use disorders (F5_5). The factor loadings and inter-factor correlations are fully detailed in **Supplementary Table 15**. Additionally, we compared our model with the previously published four-factor model by Grotzinger et al. (2022),^10^ which identified factors representing compulsive/obsessive disorders (F1_4), thought disorders (F2_4), neurodevelopmental disorders (F3_4), and internalizing disorders (F4_4). We refer to original publication^10^ for further details of the four-factor model.

For both the five-factor and four-factor models, we generated factor-specific PRSs using the GenomicSEM.^12^ We conducted multivariate genome-wide association analyses for each factor while accounting for the correlations among factors. This approach allowed us to identify genetic variants specifically associated with each latent dimension while controlling for shared genetic influences across dimensions. For each factor-specific GWAS summary statistics, we employed PRS-CS^50^ and PLINK^51^ to calculate PRSs using the same setting as the CPG model.

### Ethics Review

The *All of Us* Research Program conducted data collection under centralized Institutional Review Board (IRB) approval, with informed consent obtained from participants. This study adhered to the ethical guidelines outlined in the *All of Us* Code of Conduct. The Massachusetts General Hospital IRB determined that this study was exempt from human subjects research as it involves a secondary analysis of de-identified data from the *All of Us* Research Program.

## ACKNOWLEDGEMENT

This work was supported by grants to PHL from the U.S. National Institute of Health (R01MH119243, R01GM148494) and by MassGeneral Brigham Department of Psychiatry. We gratefully acknowledge *All of Us* participants for their contributions, without whom this research would not have been possible. We also thank the National Institutes of Health’s *All of Us* Research Program for making available the participant data release (v7) examined in this study. The *All of Us* Research Program is supported by the National Institutes of Health, Office of the Director: Regional Medical Centers: 1 OT2 OD026549; 1 OT2 OD026554; 1 OT2 OD026557; 1 OT2 OD026556; 1 OT2 OD026550; 1 OT2 OD 026552; 1 OT2 OD026553; 1 OT2 OD026548; 1 OT2 OD026551; 1 OT2 OD026555; IAA #: AOD 16037; Federally Qualified Health Centers: HHSN 263201600085U; Data and Research Center: 5 U2C OD023196; Biobank: 1 U24 OD023121; The Participant Center: U24 OD023176; Participant Technology Systems Center: 1 U24 OD023163; Communications and Engagement: 3 OT2 OD023205; 3 OT2 OD023206; and Community Partners: 1 OT2 OD025277; 3 OT2 OD025315; 1 OT2 OD025337; 1 OT2 OD025276. This study used GWAS summary statistics obtained from the 23andMe, Inc. cohort. We would like to thank the research participants and employees of 23andMe, Inc. for making this work possible. GWAS summary statistics data used in this study were also obtained from the database of Genotypes and Phenotypes (dbGaP) under accession number phs001672.v12.p1. We thank the investigators who contributed the phenotype and genotype data to dbGaP. We thank Million Veteran Program (MVP) staff, researchers, and volunteers, who have contributed to MVP, and especially participants who previously served their country in the military and now generously agreed to enroll in the study. The MVP Program is detailed in the study by Gaziano, J.M. et al.^54^ and is based on data from the Million Veteran Program, Office of Research and Development, Veterans Health Administration. The dbGaP dataset(s) used in this study were accessed with authorized approval from the National Institutes of Health (NIH). This study also used GWAS summary statistics obtained from the Psychiatric Genomics Consortium (PGC). We would like to thank the research participants and the investigators of these studies for making the data publicly available, which was essential to conducting this study possible.

## DATA AVAILABILITY

The *All of Us* Research Program data used in this study are available to authorized researchers through the *All of Us* Research website (https://allofus.nih.gov). For GWAS summary statistics from 23andMe, Inc. are available to the researchers through a separate permission process via https://research.23andme.com/dataset-access/. GWAS summary statistics from the MVP data are available on dbGaP (accession no. phs001672). For all other publicly available GWAS summary statistics, we provided the download links in the **Supplementary Table 3**.

## CODE AVAILABILITY

For all analyses, we used open-source software packages from R (v4.3) available in the *All of Us* Research Workbench (https://www.researchallofus.org/data-tools/workbench/), visualization using the R-studio desktop applications (https://posit.co/download/rstudio-desktop/), and publicly available methods GenomicSEM (https://github.com/GenomicSEM/GenomicSEM), PLINK v2.0 (https://www.cog-genomics.org/plink/2.0/), and PRS-CS (https://github.com/getian107/PRScs) as summarized in the Methods section.

